# Missense *ABI2* variants linked to a neurodevelopmental disorder with intellectual disability, epilepsy, hypoplasia of the corpus callosum, and white matter abnormalities

**DOI:** 10.1101/2025.05.19.25327541

**Authors:** Emanuela Argilli, Caleb Yang, Carolyn Le, Amanda M. Elashoff, Kendall C. Parks, Clare Bakker, Brian G. Skotko, Nancy Pinnell, Sonal Mahida, Heather Olson, Kimberly Amburgey, James J. Dowling, Louisa Kalsner, Deepa S. Rajan, Christine Munro, Christopher Barnett, Alicia B. Byrne, Eleina M. England, Erfan Aref-Eshghi, Sureni V. Mullegama, Michelle M. Morrow, Elizabeth George, Elliott H. Sherr

**Author notes:** Address correspondence to: Elliott H. Sherr, MD PhD 675 Nelson Rising Lane, 214B; and Emanuela Argilli, PhD 675 Nelson Rising Lane, 290D.

## Abstract

The Abelson-interactor 2 gene (*ABI2)* encodes a protein that functions as a regulator of Rac-dependent actin cytoskeleton dynamics, a highly coordinated structural framework essential for maintaining intracellular homeostasis and vital in processes such as cell adhesion, communication, membrane transport, migration, cell growth, and development. As a component of the Rac-1 activated WAVE regulatory complex (WRC), ABI2 initiates the actin polymerization machinery Arp2/3 to drive lamellipodia formation, and underlying key cellular processes such as axonal guidance, cell motility, and cell adhesion. Additionally, ABI2 acts as a substrate for non-receptor tyrosine kinases ABL1 and ABL2, with downstream effects controlling neuronal differentiation and migration involved in neocortical development.

Here, through exome sequencing and international collaborations, we identify eight unrelated individuals with severe neurodevelopmental delays linked to heterozygous *ABI2* missense variants, including a recurrent p.Tyr491Cys in the highly conserved SH3 domain in six individuals. These variants arose *de novo* in cases where parental testing was available, and were associated with moderate to severe motor delay, absent or delayed expressive language, intellectual disability, seizures, autistic traits, as well as macrocephaly, thinning of the corpus callosum, and remarkable white matter signal abnormalities. This report adds *ABI2* to the list of genes implicated in neurodevelopmental disorders, with an additional focus on epilepsy and brain malformations.

## Introduction

*ABI2* (GenBank: <u>NM_001375670.1</u>, MIM: <u>606442</u>) is located on cytogenetic band 2q33.2. It encodes the ABI2 protein, whose expression and cellular localization support its role in the developing nervous system. The protein acts as a target of Abl-family non-receptor tyrosine kinases ABL1 and ABL2,^1^ and is a key regulator of Rac-dependent cytoskeleton organization, involved in several signaling pathways associated with actin polymerization. A knockout *abi2* mouse model is characterized by cell migration defects in the neocortex and hippocampus, abnormal dendritic spine morphology, and density resulting in cortical and corpus callosum anomalies.^2^

ABI2 regulates the neuronal transition from multipolar to bipolar, and the interaction of growth cones with glial processes, thus playing an important role in initiating and controlling neuronal migration and proper cortical development.^3^ Numerous components of the Rac1-activated actin cytoskeleton dynamics pathway have been implicated in neurodevelopmental syndromes characterized by cognitive delays, seizures, and brain abnormalities—yet *ABI2* has not been previously associated with a human syndromic disorder. Here, we report the first cohort of individuals with heterozygous missense *ABI2* variants, including a recurrent p.Tyr491Cys in six unrelated individuals. The associated disorder is characterized by developmental and cognitive delay, epilepsy, autistic traits, and brain anomalies. While two probands with autism spectrum disorder (ASD) and two siblings with intellectual disability (ID) have been previously reported carrying monoallelic and biallelic variants in *ABI2,*^4,5^ this is the first description of a robust cohort highlighting the clinical spectrum of *ABI2*-related neurodevelopmental disorder.

## Methods

Individuals 1, 2, 3 and 5 were research participants in the ‘Disorders of Cerebral Development: A Phenotypic and Genetic Analysis Study’ at the Brain Development Research Program (BDRP) of the University of California, San Francisco (UCSF) as part of a larger cohort study of individuals with corpus callosum abnormalities. This study protocol was approved by the UCSF Institutional Review Boards (IRB), and the enrolled probands had informed consent provided by the parents/guardians.

Individuals 2, 3, 4, 6, 7, and 8 were identified through MatchMaker Exchange.^6^ Informed consent for the individuals not enrolled in the UCSF BDRP study was obtained from their respective institutions. Available medical records were collected from each research participant and reviewed by the UCSF team, including a pediatric neurologist. Full or partial de-identified MRI scans for individuals 1, 2, 3, 4, 5, and 7 were also transferred to UCSF and centrally reviewed by a pediatric neuroradiologist.^7^

Exome sequencing for Individuals 1 and 5 was performed by the Genomics Platform at the Broad Institute of MIT and Harvard. Libraries from DNA samples were created with an Illumina Nextera exome capture (37 Mb target) and sequenced (150 bp paired reads) with a mean target coverage of >80x. Bioinformatics analyses were performed according to the best practices of Genome Analysis Toolkit (GATK) HaplotypeCaller package (V.3.5). Bidirectional sequences were assembled and aligned to reference gene sequences based on human genome build GRCh38/hg38 Variant filtering was based on minor allele frequency <0.1% for autosomal dominant (AD) searches, and a predicted deleterious impact on the protein.

Individuals 2, 3, 4, 6, and 7 were tested through the clinical sequencing company GeneDx. Using genomic DNA from the proband and available relatives, the exonic regions and flanking splice junctions of the genome were captured using the IDT xGen Exome Research Panel v1.0 (Integrated DNA Technologies, Coralville, IA). Massively parallel (NextGen) sequencing was done on an Illumina system with 150bp paired-end reads. Reads were aligned to human genome build GRCh37/UCSC hg19 and analyzed for sequence variants using a custom-developed analysis tool. Additional sequencing technology and variant interpretation protocol have been previously described.^8^ We report all *ABI2* variants on the now canonical, MANE select, transcript NM_001375670, but they were originally reported on transcript NM_001282925.

## Results

From a large trio-based exome sequencing cohort of individuals with dysgenesis of the corpus callosum, we initially identified two unrelated participants with the same *de novo* variant *ABI2*:c.1472A>G (p.Tyr491Cys). Via Matchmaker Exchange, we identified six more individuals with missense variants and overlapping phenotype, including four more with the recurrent variant p.Tyr491Cys, one c.1388T>C (p.Val463Ala), and one c.1348C>T (p.Pro450Ser).

The canonical ABI2 protein sequence is 542 amino acids (NCBI Reference Sequence: NM_001375670.1, Figure 1A). Amino acids 93-156 form the Abl-interactor homeo-domain homologous domain (Abi_HHR), which contributes to its cellular localization in the lamellipodium.^9^ Amino acids 481-541 form the Src-homology 3 (SH3_Abi2) domain. ABI2 interacts via reciprocal phosphorylation of its SH3 domain with non-receptor tyrosine kinases ABL1 and ABL2.^1,10^

**Figure 1.**
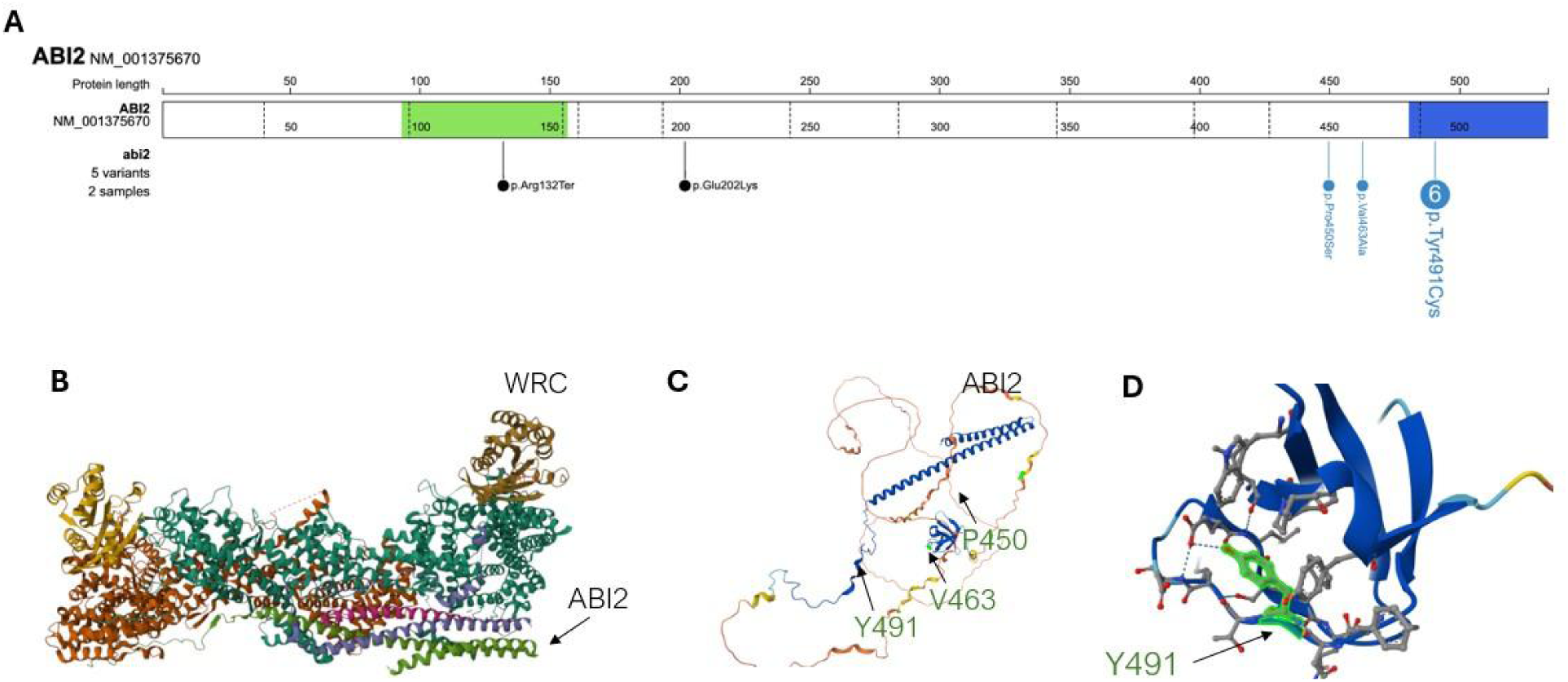
Localization of ABI2 variants. (A) Schematic representation of the variants in ABI2 reported in our cohort (in blue) and previously reported (in black), created with ProteinPaint. (B) 3D model of the WRC assembly with ABI2 coiled-coil domain indicated by the arrow. (C) 3D model of the ABI2 protein with the location and amino acids affected by variants in this study, and (D) a close-up of the SH3 domain with site Y491 (UNP:F8WAL6).

The *ABI2* p.Tyr491Cys variant carried by six unrelated individuals described here was confirmed *de novo* when parental DNA was available for testing (4/6). This variant is located within the SH3 domain (Figure 1A), is absent from the Genome Aggregation Database (gnomAD v4), and is predicted unanimously to have a deleterious effect on protein function by in-silico scores (CADD: 32, REVEL: 0.61, MutPred: 0.87, MPC: 1.88, Eigen: 7.50, MutTaster: disease causing, PolyPhen: 1.000).^11^ Tyrosine is an aromatic amino acid with a hydroxyl group, while cysteine has a more hydrophilic sulfhydryl group. The non-conservative cysteine to tyrosine substitution can alter the protein’s overall hydrophobicity, potentially affecting its structure, folding, stability, interactions, and cellular localization (Figure 1B, 1C, 1D).

The *ABI2* p.Val463Ala variant was observed *de novo* in individual 7. It is absent from gnomAD databases, has a CADD score of 25.5, a REVEL score of 0.24, and is located between the disordered region and the SH3 domain. A valine to alanine substitution might have a less drastic effect on protein structure. However, it can still have significant consequences. Alanine is a smaller, more flexible, and less hydrophobic amino acid, with a methyl group as its side chain instead of a branched-chain.

The *ABI2* p.Pro450Ser, present in Individual 8 with unknown origin, is located in the disordered region, has a CADD score of 23.6, a REVEL score of 0.28, and the gnomAD allele frequency is 6.20e-7. A proline to serine substitution can disrupt the protein’s folding and stability. Proline has a unique three-carbon side chain that cyclizes with the amino group, creating a pyrrolidine ring. This rigid structure may disrupt regular secondary structures like alpha-helices and beta-sheets. Serine, on the other hand, is smaller and more flexible, allowing for greater conformational freedom.

Table 1 compiles genetic and clinical data for individuals with germline *ABI2* variants. The cohort includes eight individuals—four males and four females. One was a fetus, while the others ranged in age from 0 to 30 years. All, except the fetus, were born at term without prenatal concerns, aside from one case of decreased fetal movements requiring maternal bed rest later in pregnancy.

**Table 1.** Molecular and phenotypic data for individuals with *ABI2* variants.

|  | Individual 1 | Individual 2 | Individual 3 | Individual 4 | Individual 5 | Individual 6 | Individual 7 | Individual 8 |
| --- | --- | --- | --- | --- | --- | --- | --- | --- |
| Genetic |  |  |  |  |  |  |  |  |
| *ABI2* variant (hg38) | 2:20342719<br>5 A>G | 2:20342719<br>5 A>G | 2:20342719<br>5 A>G | 2:20342719<br>5 A>G | 2:20342719<br>5 A>G | 2:20342719<br>5 A>G | 2:203417016<br>T>C | 2:203416976<br>C>T |
| Transcript<br>NM_001282925 | c.1385 A>G,<br>p.Ty462Cys | c.1385 A>G,<br>p.Ty462Cys | c.1385 A>G,<br>p.Ty462Cys | c.1385 A>G,<br>p.Y462C | c.1385 A>G,<br>p.Ty462Cys | c.1385 A>G,<br>p.Ty462Cys | c.1301T>C,<br>p.Val434Ala | c.1261C>T,<br>p.Pro421Ser |
| Canonical<br>Transcript<br>NM_001375670 | c.1472A>G,<br>p.Tyr491Cys | c.1472A>G,<br>p.Tyr491Cys | c.1472A>G,<br>p.Tyr491Cys | c.1472A>G,<br>p.Tyr491Cys | c.1472A>G,<br>p.Tyr491Cys | c.1472A>G,<br>p.Tyr491Cys | c.1388T>C,<br>p.Val463Ala | c.1348C>T,<br>p.Pro450Ser |
| Inheritance | de novo | unknown | unknown | de novo | de novo | de novo | de novo | unknown |
| GnomAD v4<br>frequency | 0 | 0 | 0 | 0 | 0 | 0 | 0 | 1.24E-06 |
| Developmental<br>features |  |  |  |  |  |  |  |  |
| Sex | female | female | female | male | female | male | male | male |
| Born at term | + | + | N/A | N/A | N/A | + | + | - (pregnancy<br>termination) |
| OFC (SD) | +2.7 | >2 | +4.2 | +4.04 | N/A | +3.11 | +2.5 | N/A |
| Developmental<br>delay | +(moderate) | +(severe) | +(severe w/<br>regression) | +(severe) | +(severe) | +(severe) | +(moderate<br>w/<br>regression) | N/A |
| Motor delay | +(moderate) | +(severe) | +(w/<br>regression) | + | + | + | + | N/A |
| Speech delay | + | +(words) | +(words) | +(non-verbal) | +(non-verbal) | +(non-verbal) | +(moderate<br>w/<br>regression) | N/A |
| Neurological<br>features |  |  |  |  |  |  |  |  |
| Intellectual<br>disability | + | + | +(moderate-severe) | +(severe) | +(severe) | +(moderate-severe) | + | N/A |
| Epilepsy (onset;<br>type) | +( >2 years;<br>absence<br>seizures,<br>negative<br>EEG) | +( <2 year;<br>infantile<br>spasm,<br>generalized<br>tonic-clonic<br>and<br>absence<br>seizures,<br>intractable) | +( <2 years;<br>complex<br>partial,<br>generalized<br>tonic-clonic<br>seizures) | +( >2 years;<br>complex<br>partial,<br>multifocal,<br>absence<br>seizures) | +( <2 year;<br>severe) | +( <2 years;<br>complex<br>partial,<br>generalized<br>tonic-clonic<br>seizures) | - | N/A |
| ASD traits | + | +(ASD) | N/A | +(ASD) | N/A | + | +(ASD) | N/A |
| ADHD | + | + | N/A | + | N/A | N/A | - | N/A |
| Tone/gait<br>anomalies | +(hypotonia) | +(hypotonia,<br>unsteady<br>gait) | +(hypotonia,<br>unsteady<br>gait) | +(hypotonia) | N/A | +(hypotonia) | - | N/A |
| Feeding/swallowing<br>problems | - | - | + | - | N/A | + | - | N/A |
| Brain MRI |  |  |  |  |  |  |  |  |
| Hypoplastic<br>corpus callosum | + | + | + | + | + | + | - | +(short and<br>hypoplastic) |
| Hypoplastic<br>anterior<br>commissure | + | + | + | + | + | N/A | - | N/A |
| Enlarged ventricles | - | + (mild) | + (frontal horns, mild) | + (frontal horns, mild) | + (mild) | + | - | - |
| Persistent cavum septum pellucidum | + | + | + | + | + | N/A | - | N/A |
| Low WM volume | + | + | + | + | + | + | - | N/A |
| Abnormal myelination | - | - | - | - | - | N/A | - | N/A |
| Periventricular T2-FLAIR hyperintensity in WM | + (P>A) | + (P>A, stable over time, or improving) | + (P>A, stable over time) | + (P>A) | + (P>A) | N/A | - | N/A |
| Other | +(left cerebellar medullary angle cyst) | + (findings of hippocampal sclerosis; small cystic spaces or enlarged perivascular spaces in WM) | - | - | - | + (findings of hippocampal sclerosis) | - | - |
| Clinical features |  |  |  |  |  |  |  |  |
| Vision/oculomotor | - | + (infantile esotropia with L amblyopia) | + (cortical visual impairment) | - | N/A | - | - | N/A |
| Cardiovascular | - | - | + (dilated coronary arteries in infancy, resolved) | - | N/A | - | - | + (vascular ring) |
| Dysmorphic features | - | + (mild cavus deformities) | + (long face, bitemporal hollowing, prominent forehead, short nose, anteverted nares, broad nasal root, thick lips, broad great toes) | - | N/A | + (prominent forehead, flat nasal bridge, thin and tented upper lip) | + (prominent forehead, nonspecific facial dysmorphism) | + (mild microglossia; elongated hand digits) |
| Other |  | Heel cord tightness; chronic idiopathic constipation | hepatomegaly in infancy, resolved | Likely arachnoid cyst in the mid-thoracic level | N/A |  |  |  |

Abbreviations: + = Present, - = Absent, OFC = Occipitofrontal Circumference, SD = Standard Deviation, WNL = Within Normal Limits, ASD = Autism Spectrum Disorder, ADHD =
Attention-Deficit/Hyperactivity Disorder, P = posterior, A = Anterior, WM = White Matter, VUS = Variant of Uncertain Significance

In the individuals available for postnatal evaluation, development was characterized by moderate to severe delays across all domains (7/7), including gross and fine motor skills, speech and language, cognition, and social interactions. Walking was achieved between the ages of 2 and 9 years, with some requiring physical support. Generalized hypotonia was reported in all individuals with available data (5/5). Individual 3 presented with mixed tone (including hypotonia and spasticity), and crouched gait. Among six individuals aged 5 years and older, three (3/6) were non-verbal. Within the subset of individuals aged 10 years or older, one (1/4) remained non-verbal, two (2/4) used single words, and one (1/4) developed the ability to form full sentences. Evidence of regression was observed in two individuals. Moderate to severe ID was present in all individuals (7/7). Autism spectrum disorder (ASD) characteristics were observed in all the individuals evaluated (5/5), with a formal ASD diagnosis in three (3/5).

Individuals 1 through 6 carrying the p.Tyr491Cys variant all had seizures. The onset of epilepsy was before the age of 2 years in 5/6 individuals and characterized by complex partial and generalized tonic-clonic seizures, with an abnormal electroencephalogram (EEG). In one individual, seizure onset was perinatal with infantile spasms; another had only one febrile seizure before the age of 2 years and a series of absence and tonic-clonic seizures after the age of 5 years, with a normal continuous video EEG with no epileptiform activity or focal abnormalities detected.

We highlight striking similarities on brain MRI (Figure 2). All six individuals with the p.Tyr491Cys variant had hypoplastic corpus callosum, characterized by moderate to severe thinning of all regions. When assessed, the anterior commissure was also hypoplastic (5/5). Myelination was normal, but there were significant white matter anomalies. Hyperintense signals in the periventricular white matter T2-weighted Fluid-Attenuated Inversion Recovery (T2-FLAIR) sequences were observed in 6/6 scans, together with mildly decreased white matter volume. A persistent cavum septum pellucidum was present in 5/5. The ventricles were slightly enlarged and dysmorphic in 5/6, particularly the frontal horns. Two individuals had findings consistent with mesial temporal (or hippocampal) sclerosis.

**Figure 2.**
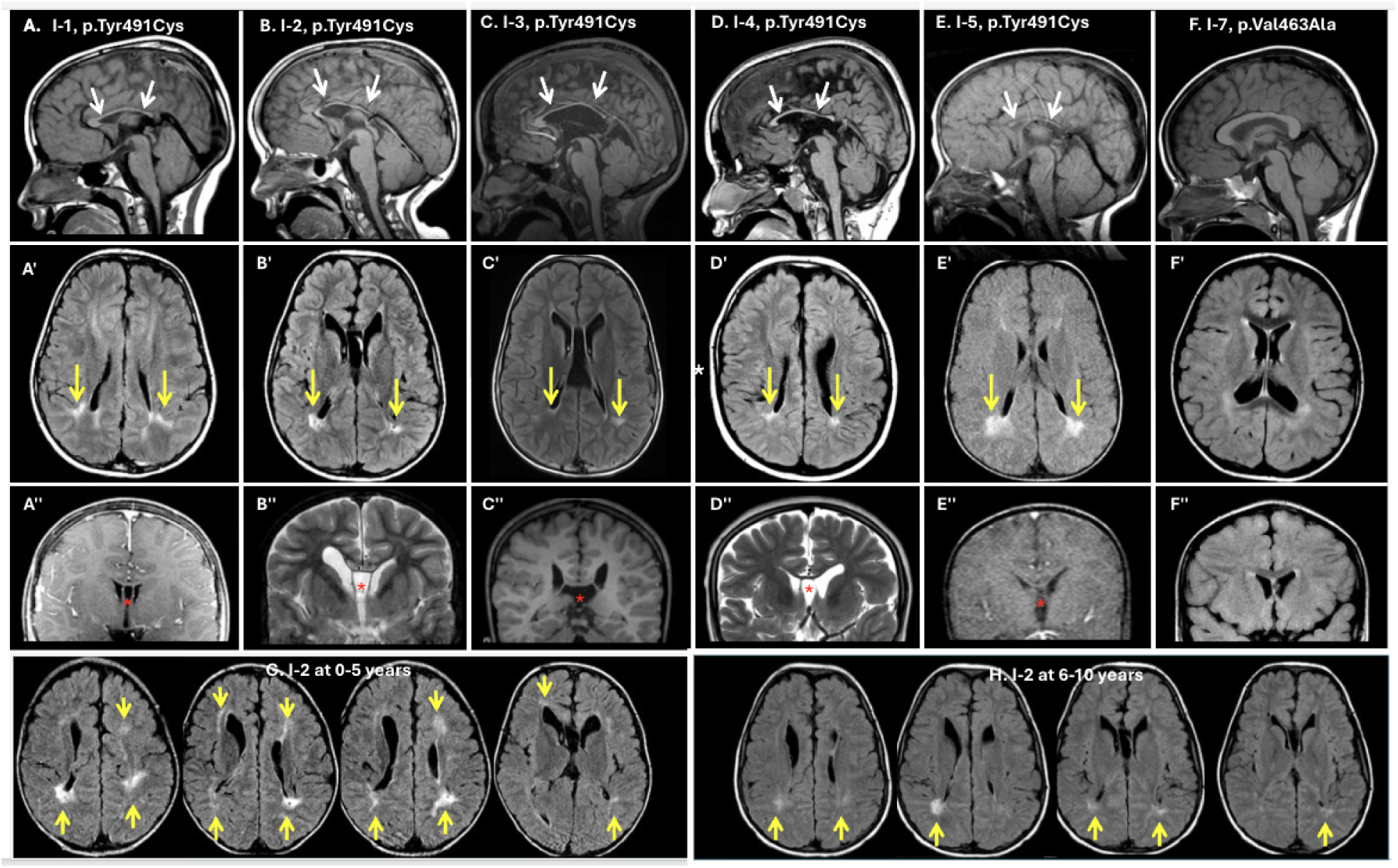
Neuroradiologic features of individuals with *ABI2* variants. Representative brain MR images highlight remarkably similar features in individuals with the p.Tyr419Cys variant. Top row: Sagittal T1 images showing hypoplasia of the corpus callosum (white arrow) in I-1 (A), I-2 (B), I-3 (C), I-4 (D), I-5 (E) with the p.Tyr419Cys variant. I-7 (F) with the p.Val463Ala variant showed a normal corpus callosum. Middle row: Axial T2-FLAIR images in the same individuals as above panel showed patchy signal abnormality and decreased volume of the periventricular white matter (yellow arrows) as well as dysmorphic, mildly enlarged ventricles (A’, B’, C’, D’, E’) except I-7 (F’). Third row: Coronal T1 or T2 images showed a persistent cavum septum pellucidum (red asterisk) in all (A’’, B’’, C’’, D’’, E’’) except I-7 (F’’). Bottom row: Repeated scans at the age of 0-5 years (G) and 6-10 years (H) in Individual 2 showed mild improvement in the white matter anomalies over time, particularly anteriorly.

Individual 2 had repeated brain MRIs available for comparison between infancy and 9 years. While white matter signal abnormalities remained present, the areas of hyperintensity were less prominent at 6-10 years compared to 0-5 years (Figure 2. J, H). After the age of 2 years, small cystic spaces or dilated perivascular spaces were observed in the periventricular white matter. Ictal and interictal Single-Photon Emission Computed Tomography (SPECT) showed mild interictal hypoperfusion in the left temporal lobe, and bilateral regions of higher ictal than interictal perfusion, particularly in the antero-inferior right temporal lobe, suggesting a potential focus for seizure.

Individual 3 also had repeated MRIs between infancy and 9 years which showed stable ventricles size and white matter abnormalities over time. Individual 7 (p.Val463Ala) had a normal MRI; mild peritrigonal white matter hyperintensity on T2-FLAIR was within normal limits (Figure 2, F’). Individual 8 (p.Pro450Ser) had a fetal brain MRI in the second trimester that revealed a short and hypoplastic corpus callosum. No gross abnormalities were noted in the white matter.

All individuals assessed (6/8) had an enlarged occipital-frontal circumference (OFC), ranging from +2.5 SD up to +4.2 SD, and macrocephaly present at birth in 2/4 individuals. Craniofacial dysmorphisms beyond macrocephaly were observed, including prominent forehead (3/6), flat or broad nasal bridge (2/6), and the following features each present in 1 of 6 individuals: short nose, anteverted nares, thin and tented upper lip, thick lips, low anterior hairline, bitemporal hollowing, and long face. Other anomalies included long fingers, broad great toes, and mild pes cavus.

Two individuals had cardiovascular anomalies. One had dilated left anterior descending and proximal right coronary arteries in infancy (resolved), and one has a vascular ring on fetal scan. Ophthalmological issues were seen occasionally (2/6) and included one individual with infantile esotropia with left amblyopia, and one with cortical visual impairment.

## Discussion

ABI2 plays a key role in actin-dependent cytoskeleton organization and dynamics, a well-known signaling pathway crucial to the developing brain. Prior to this manuscript, ABI2 had yet to be strongly linked to a neurodevelopmental brain disorder. In this study, we provide such evidence detailing the findings of a cohort of eight individuals with heterozygous *ABI2* variants who also had moderate to severe developmental and cognitive delay, seizures, and characteristic brain abnormalities.

The neurodevelopmental phenotype was characterized by severely delayed walking, unsteady gait, generalized hypotonia, absent or delayed expressive language, intellectual disability, characteristics of autism spectrum disorder, and macrocephaly. Individuals with the recurrent p.Tyr491Cys variant (6/8) had epilepsy of variable semiology. They also shared remarkable similarities on brain imaging. Specifically, all had a diffusely thin corpus callosum, a small anterior commissure, periventricular white matter signal abnormality, mildly enlarged and dysmorphic ventricles, low white matter volume, and a persistent cavum septum pellucidum. Only one individual had a normal brain and callosal phenotype and carried the variant p.Val463Ala. We hypothesized that missense variants in the SH3 region or near its boundary compromise the protein subcellular location or its ability to bind substrates,^12,13^ but more conservative amino acid substitutions, such as this valine to alanine, might be less disruptive.

The human ABI2 protein is expressed in most tissues but is also most abundant in the central nervous system (CNS). It is a key regulator of actin cytoskeleton dynamics, as a component of the WAVE regulatory complex (WRC), and via interaction with non-receptor tyrosine kinase ABL1. ABI2 positively regulates ABL1-mediated phosphorylation of ENAH,^1,10,14^ required for proper polymerization of nucleated actin filaments at the leading edge, influencing cell growth, differentiation, and axon guidance, essential events of CNS development.^15,16^

The WRC consists of a Sra1/CYFIP1-Nap1/NCKAP1 heterodimer and a WAVE1-ABI2-BRICK1 heterotrimer. Expression of these proteins in human fetal tissues suggests an important role in fetal development.^17^ WAVE1 has a verprolin homology-central-acidic (VCA) domain that interacts with CYFIP1 in its inhibited state.^18^ RAC1, a Rho family GTPase, binds to the VCA domain to activate WAVE1 and promote Arp2/3-mediated branched actin filament formation.^19–23^ Downstream effects include the formation of lamellipodia, adherens junctions, neurite growth cones, and dendritic spines, as well as axon branching, axon projection, pathfinding, and sorting.^24–27^ *WAVE1* (*WASL1*) and *RAC1* variants both negatively affect the organization of actin bundles and disrupt the formation of lamellipodia in human fibroblast models from affected individuals.

In mice, Abi2 expression is enriched in the developing brain and spinal cord, suggesting a role in cortical development. In the fetal brain, Abi2 localizes to post-migratory, differentiated neurons while in postnatal brains, it is prominent in projection neuron populations and regions that exhibit synaptic plasticity. Within the neocortex, Abi2 is abundant in the cortical plate’s differentiated neurons and glia, rather than in the intermediate or ventricular zones. Peripheral expression of Abi2 is also elevated in the dorsal root ganglia.^28^

Notably, corpus callosum anomalies have been shown in Abi2-deficient mice. These mice exhibited impaired neuronal migration in the neocortex and hippocampus, with aberrant localization of cortical cells into the corpus callosum, regions of low cellularity surrounded by misoriented neurons in deep layers of the motor and visual cortices. *Abi2* knockout mice also exhibited impaired formation of adherens junctions and abnormalities in dendritic spine morphology.^2^ Similar anomalies have been observed in vitro following the expression of dominant-negative Rac. It has been suggested that the loss of Abi2 disrupts Rac-dependent pathways regulating actin dynamics, ultimately leading to deficits in spine morphology and density.^29^ Additionally, the Grove et al. study^2^ showed defective orientation and migration of secondary lens fibers into the eye. One individual in this cohort had cortical visual impairment.

Many syndromic autosomal dominant neurodevelopmental disorders have been strongly linked to missense variants affecting proteins involved in actin-mediated cytoskeleton regulation, including many of the WRC five core proteins—WAVE1, Nap1, Sra1, ABI2, and HSPC3000. *WASL1* (WAVE1) variants have been associated with ID, seizures, and white matter anomalies.^26,30^ *NCKAP1* (Nap1) variants were linked to ID and ASD,^31^ and *CYFIP2* (Sra1) variants to ID, epilepsy, ASD, and white matter anomalies.^32^ Pathogenic *RAC1* and *CDC42* variants, affecting key proteins that function as activators of the WRC, cause ID, seizures, reciprocally macrocephaly and microcephaly, corpus callosum hypoplasia, and white matter signal abnormalities.^25,33^ Studies indicate that CDK5, along with constitutively active CK2 kinases, phosphorylates the WRC, preventing it from interacting with the Arp2/3 complex, thus disturbing actin polymerization.^34,35^ *CDK5* variants have a provisional association with a recessive disorder with a lack of psychomotor development, seizures, and complex brain malformations.^36^ The *ABI2-*associated syndrome we described shares many clinical features attributed to a disrupted actin polymerization cytoskeleton pathway.

Heterozygous variants in *ABL1,* binding partner for ABI2, are associated with a disorder involving congenital heart defects, facial dysmorphisms, and failure to thrive. Corpus callosum hypoplasia was observed in one individual.^37^ In the ABI2 cohort, individual 3 had dilated proximal coronary arteries in infancy, and individual 8 had fetal cardiac anomalies.

Before our study, the evidence linking *ABI2* variants to a human neurodevelopmental disorder was minimal. In one publication, two unrelated probands from a cohort of 2,500 individuals with ASD, one with normal intelligence and one with borderline ID, carried *de novo* variants in *ABI2:*p.Glu202Lys and *ABI2*:p.Arg373Cys, respectively.^4^ It was speculated that disruptions of Contactins-NYAPs-WAVE1 signaling is an important pathogenesis pathway in ASD etiology.^38^ In a second publication, a homozygous nonsense variant *ABI2:*p.Arg132* was found in two siblings from a cohort of 192 individuals with ID and consanguinity.^5^ Of note, in this family, two out of three additional siblings had ID, and a heterozygous *ABI2*:p.Arg132* variant inherited from one unaffected parent.

Here, we present a robust cohort of individuals with *ABI2* heterozygous variants that begins to define the details of the genetic, clinical, and imaging phenotypic spectrum of a novel human neurodevelopmental disorder. Six individuals carried the identical variant p.Tyr419Cys. This change is the highly conserved SH3 region of the protein and is unanimously predicted to be deleterious. The overlap between clinical and neuroradiological phenotypes of individuals who harbor this variant is very robust, strongly suggesting this is a hotspot for pathogenic genetic variations. Because ABI2 effectively regulates neuronal migration in the neocortex via interaction with the Rac-pathway, and Abi2-lacking mice have impaired migration affecting the corpus callosum, we suggest that variants in *ABI2* (particularly, the recurrent Tyr491Cys) contribute to the brain abnormalities observed, including corpus callosum hypoplasia. More research on this variant is needed to determine if the observed features are primarily linked to disruption of the ABL1 or the WAVE-Rac pathway, and its impact on neuronal morphology and differentiation in neocortical development.

In conclusion, through this multicenter collaboration study, we expand the list of actin-regulatory-pathway genes linked to neurodevelopmental disorders with associated brain abnormalities. Based on the present cohort, *ABI2*’s addition to the gene testing panel for intellectual disability and epilepsy is warranted; although even more appropriate would be a broader test, such as exome/genome sequencing, which is recommended by the American College of Medical Genetics and Genomics (ACMG) for patients with congenital anomalies or ID.^39^ This approach would identify *ABI2* as well as other actin-regulatory pathway genes that may be recognized in the future.

## Data Availability

All data produced in the present study are available upon reasonable request to the authors

## Acknowledgements

We thank the families who participated in this study. This work was supported by NIH grant R01NS058721. Sequencing and analysis of Individuals 1 and 5 were provided by the Broad Center for Mendelian Genomics, funded by the National Human Genome Research Institute grants UM1HG008900, U01HG0011755, and R01HG009141.

## Web Resources

ExAC Browser, http://exac.broadinstitute.org/

Genome Aggregation Database (gnomAD), https://gnomad.broadinstitute.org Matchmaker Exchange, http://www.matchmakerexchange.org/

Online Mendelian Inheritance in Man, OMIM®, https://omim.org/ GeneCards, https://www.genecards.org/

The Human Protein Atlas, https://www.proteinatlas.org/ AlphaFold Protein Structure Database, https://alphafold.ebi.ac.uk/ CADD, https://cadd.gs.washington.edu/

ClinVar, https://www.ncbi.nlm.nih.gov/clinvar DECIPHER, https://decipher.sanger.ac.uk/ GenBank, https://www.ncbi.nlm.nih.gov/genbank/

GeneDx ClinVar submission, http://www.ncbi.nlm.nih.gov/clinvar/submitters/26957/ Uniprot, https://www.uniprot.org/

